# Delirium in a Young Predominantly Hispanic Population with COVID-19

**DOI:** 10.1101/2023.01.22.23284879

**Authors:** Kelsey C. Talkington, Luis A. Alvarado, Christopher A. Castaneda, Silvina B. Tonarelli

## Abstract

**Purpose:** To study a primarily Hispanic population of adults younger than 65 to determine if COVID-19 patients with a concurrent delirium diagnosis had worse clinical outcomes in terms of hospital stay, ventilation and mortality, than those without a delirium diagnosis.

**Methods:** After approval by the appropriate Institutional Review Board, a retrospective cohort study was performed looking at demographics, vital statistics, and clinical outcomes of patients aged 18-65 admitted to a hospital in the United States – Mexico border region with COVID-19 between March 1 and June 30, 2020. Data were analyzed using Fisher’s exact test, or an unpaired *t*-test where appropriate, and a univariate analysis was performed to establish relative risk. Confidence intervals were set at 95% and *p* values ≤0.05 were considered significant.

**Results:** 133 patients with confirmed COVID-19 diagnoses (58% men, 92% Hispanic) were included. Mean age was 50.5 with a standard deviation of 11.7 years (range 20-65 years). The prevalence of delirium was 6%. Fifty percent of delirium patients died during hospitalization compared to 15% of patients without delirium. Patients with delirium were found to spend more days hospitalized, in the intensive care unit, and intubated than their counterparts without delirium. Delirium was associated with increased risk of being placed on mechanical ventilation (RR 3.91, 95% CI 1.46—10.41, *p* value 0.006).

**Conclusions:** Delirium was associated with worse COVID-19 outcomes independent of age. COVID-19 patients need to be actively assessed for signs of delirium and appropriate precautionary measures should be implemented. Proper documentation of delirium is key to continue learning about the incidence of delirium in COVID-19 patients.

---

Severe acute respiratory syndrome coronavirus 2 (SARS-Cov-2) causes coronavirus disease 2019 (COVID-19), which the WHO declared a pandemic on March 11, 2020.^1^ COVID-19 is a disease with a highly variable acute presentation that may include delirium.^2,3^ Younger COVID-19 patients, those 65 and under, may present with different symptoms than those over 65.^4–6^ Delirium is more prevalent in older populations with existing cognitive issues.^7,8^ Delirium is a mental disorder described in the Diagnostic and Statistical Manual of Mental Disorders, 5^th^ ed. (DSM-5) as defined in part by a disturbance in attention and awareness which can develop over a short period of time.^9^

Delirium is prevalent yet underdiagnosed in the Emergency Department (ED), Intensive Care Unit (ICU) and on medical floors, and has been linked to adverse clinical outcomes like increased mortality and increased length of hospital stay in ICU and non-ICU settings.^8, 10–13^ Patients under 65 years old have demonstrated prevalent delirium in ICU and intermediate care units (IMC) prior to the appearance of COVID-19.^13^ In patients with COVID-19, delirium is being recognized as a common complication and, potentially, a symptom in atypical presentations of the disease^14,15^ A study of 707 COVID-19 patients in a Brazilian teaching hospital indicated that in older adults with COVID-19, a diagnosis of delirium was associated with increased adverse health outcomes while hospitalized.^16^

Studies indicate that in older adults preexisting psychiatric diagnoses are associated with an increased rate of delirium with COVID-19.^3,15^ Other studies have excluded those with psychiatric diagnoses and existing brain problems, potentially leading to underestimation of rates of delirium in COVID-19 patients and ignoring a potentially significant comorbidity for adverse COVID-19 outcomes.^17^

There have been concerns of a dearth of research regarding COVID-19 presentation and outcomes in ethnic minority populations in the United States.^18^ Studies have since found COVID-19 to disproportionately affect minorities such as African Americans, Native Americans and Hispanic communities.^19^ Our study was done in a county teaching hospital which serves the predominantly Hispanic population (83%) of the US/Mexico border city of El Paso and so is an excellent setting for studying COVID-19 presentations and outcomes in this ethnic group.^20^

Thus, we hypothesized that of COVID-19 patients admitted to this hospital, those with concurrent diagnoses of COVID-19 and delirium would have more adverse clinical outcomes, including an elevated mortality rate, than those without a delirium diagnosis, regardless of age. We explored associations between adverse clinical outcomes such as intubation and the presence of delirium. We studied associations of mortality, days alive after hospital admission, and days hospitalized in COVID-19 patients with concurrent delirium diagnoses, and the relationship of existing mental health diagnosis with development of delirium in COVID-19 patients.

## Methods

A retrospective cohort study was performed looking at patients admitted to a county medical teaching hospital on the United States (US)—Mexico border (the Hospital) with a COVID-19 diagnosis. The study was deemed exempt and approved by the relevant Institutional Review Board (IRB) and granted waiver of informed consent because data were deidentified. The hospital database (CERNER) was reviewed to identify records of all patients aged 18 to 65 years with a COVID-19 diagnosis between March 1, 2020 and June 30, 2020, and those patients admitted to the Hospital were enrolled in the study. Three hundred seventy three patient records were identified and 133 were found to meet the selection criteria, resulting a sample size of 133. Patient demographics (age, gender, race, ethnicity), initial chief complaints at the Emergency Department (ED) (abdominal pain, chills, rash, fatigue, fever, headache, muscle pain, new or worsening cough, persistent cough with blood or mucous, shortness of breath, diarrhea, vomiting, weakness/numbness), health measures (temperature, heart rate, respiration rate, systolic blood pressure, diastolic blood pressure, SpO_2_, oxygen therapy), hospitalization course (COVID-19 diagnosis, delirium diagnosis, days with delirium, length of hospital stay, outcome of hospitalization [discharge or death], admission date, date of discharge or death, mortality status 30 days after admission, ICU admission, length of ICU stay, days on mechanical ventilation), and preexisting conditions (diabetes, hypertension, body mass index (BMI), depression, anxiety, schizophrenia, bipolar disorder, other mental health diagnoses [not including substance use disorders], cognitive deficits, history of substance use, substance used) were collected.

These data were entered into the affiliated institution’s Research Electronic Data Capture (REDCap) for data collection, deidentification, transmission and storage. REDCap is a secure web platform for building and managing online databases and is Health Insurance Portability and Accountability Act (HIPPA) compliant. All study data was entered via a password protected REDCap database website unique to this study.

### Statistical analysis

Quantitative variables were summarized using mean, standard deviation, median, and interquartile range. Categorical variables were summarized using frequency and percentages. Patients were retrospectively acquired who were admitted to the Hospital between the dates of March 1 – June 30, 2020. Patients who had delirium versus those who did not were compared using a Fisher’s exact test or an unpaired t-test. Each patient was followed after hospital admission to determine 30-day mortality status. A univariate relative risk model was used to determine the risk of delirium within the COVID-19 population (hospitalized) with respect to their ventilation status. Relative risk (RR), 95% confidence intervals (CI), and *p*-values were used to describe the model *p*-values ≤ 5% are considered statistically significant results. All analyses and data management were conducted using Stata V15.1 by the Biostatistics and Epidemiology Consulting Lab (BECL).

## Results

Three hundred seventy-three patient records were reviewed; of those a total of 133 patients met the selection criteria of COVID-19 diagnosis and admission to the Hospital and were included in the study. Of those 133 patients, 77 (58%) were male, 104 (78%) were White, 2 (1.5%) were Black or African American, 25 (19%) identified as Other. Of these patients, 120 (92%) identified their ethnicity as Hispanic. The mean (SD) age of the patients was 50.5 (11.7) years.

Symptom prevalence at initial presentation to the ED included shortness of breath (103 patients [78%]), new or worsening cough (94 patients [71%]), and fever (87 patients [66%]). Mean patient vital signs on initial presentation to the ED including temperature (°C), heart rate, respiratory rate, and SpO2 (87.5% [10.5%]) are shown in **Table 1**. Preexisting conditions included diabetes (51 patients [38%]), hypertension (70 patients [53%]), psychiatric diagnoses (25 patients [19%]), cognitive deficits (6 patients [5%]), and history of substance use (30 patients [23%]). Cognitive deficits were related to previous cerebrovascular accident and/or other causes of dementia. History of substance use included use of legal substances like tobacco and ethanol as well as illicit ones such as cocaine, marijuana, and methamphetamines. There was only one significant association across presence of any psychiatric disorder: lower diastolic blood pressure. Hospitalization outcomes including length of stay (mean 8.1, SD [7.5]), presence of delirium (8 patients [6%]), days with delirium (0.2 [1.4]), and mortality (24 patients [18%]) are captured in **Table 2**.

**Table 1.** Vital Signs at Presentation

| **Vital Sign** | **Mean (SD)** |
| --- | --- |
| **Temperature (°C)** | 37.7 (1.0) |
| **Heart Rate (beats/minute)** | 101.7 (19.5) |
| **Respiratory Rate (breaths/minute)** | 19.8 (4.2) |
| **Systolic Blood Pressure** | 130.6 (20.5) |
| **Diastolic Blood Pressure** | 77.5 (11.4) |
| **SpO<sub>2</sub> (%)** | 87.5 (10.5) |
| **Body Mass Index (kg/m<sup>2</sup>)** | 31.1 (5.6) |

**Table 2.** Hospitalization outcomes data

| **Measure** | **Patients No. (%)** |
| --- | --- |
| **Length of Hospital Stay (days), mean (SD)** | 8.1 (7.5) |
| **Days with Delirium, mean (SD)** | 0.2 (1.4) |
| **Length of ICU Stay, median (IQR)** | 0.0 (0.0, 3.0) |
| **Days on Mechanical Ventilation, median (IQR)** | 0.0 (0.0, 0.0) |
| **Mechanical Ventilation** | 25 (19) |
| **Delirium** | 8 (6) |
| **Mortality Status 30 Days from Admission** |  |
| **Alive** | 109 (82) |
| **Deceased** | 24 (18) |

The delirium patient group had a mortality rate of 50%, significantly higher than the 14.63% mortality rate for the patients without delirium (*p* = 0.027). Patients with delirium had longer hospital stays (18.0 [11.0]) and longer ICU stays (median 11.0, IQR [2.5, 23.0]) than their non-delirious counterparts whose hospital stays (7.5 [6.8]) and ICU stays (0.0 [0.0, 2.0]) were significantly shorter (*p* = <0.001). Additionally, a greater fraction of the patients with delirium were placed on mechanical ventilation, 62.5%, compared with their non-delirious counterparts, 16% (*p* = 0.006). These patients spent more days on mechanical ventilation (8.5 [0.0, 16.5]) than the patients without delirium (0.0 [0.0, 0.0], *p* = <0.001).

Male gender was significantly associated with the presence of delirium: 100% of delirium patients were male while 55.2% of patients without delirium were male (*p* = 0.021). In terms of initial presentation to the ED, patients without delirium had a lower SpO_2_, 88.3% (8.1%), than the patients with delirium, 74.5% (26.7%) (*p* = <0.001). More patients with delirium, 50%, were on O_2_ therapy at presentation compared with a rate of 9.6% in their counterparts without delirium (*p* = <0.001). Preexisting conditions significantly associated with the presence of delirium were cognitive deficits (37.5%) and history of substance use (75%).

In terms of hospitalization, those who were deceased 30 days after admission had longer hospital stays (12.2 [9.1] versus 7.3 [6.8], *p* = 0.003) and ICU stays (9.0 [5.0, 14.5] versus 0.0 [0.0, 0.0], *p* = <0.001). Significantly more had delirium (16.67% versus 3.67%, *p* = 0.035), and were placed on mechanical ventilation (87.50% versus 3.67%, *p* = <0.001), and those who were intubated spent more days on mechanical ventilation (5.5 [2.5, 11.5] versus 0.0 [0.0, 0.0], *p* = <0.001). Regarding presentation, those who were deceased 30 days from admission were older (55 ± 8.8 years compared to 49.5 ± 12.1 years, *p* = 0.036), male (79.17% versus 53.21%, *p* = <0.001), had lower SpO_2_ (77.8% [18.8%] versus 89.6% [5.9%], *p* = <0.001) and a higher respiratory rate (21.6 [7.2] versus 19.4 [3.1], *p* = 0.021) at presentation.

Twenty percent of patients who were mechanically ventilated developed delirium compared with a rate of 2.78% in their non-intubated counterparts (*p* = 0.006). In terms of initial presentation to the ED, patients placed on mechanical ventilation had lower SpO_2_ (75.7% [18%]) and higher respiratory rate (21.7 [7.2]) than their non-intubated counterparts who had SpO_2_ of 90.2% [4.9%] (*p* = <0.001) and respiration rate of 19.4 [3.0] (*p* = 0.012). Additionally, more were on O_2_ therapy: 24% versus 9.26% in their non-intubated counterparts (*p* = 0.006). No vomiting was seen in those who were intubated (100% versus 79.63%, *p* = 0.013).

Four characteristics were determined via relative risk assessment to be significant contributors to being placed on mechanical ventilation. Increased respiratory rate at presentation increased risk of intubation (RR 1.08, 95% CI 1.01—1.16, *p* = 0.022). Higher SpO_2_ at presentation decreased risk of intubation (RR 0.96, 95% CI 0.95—0.97, *p* = <0.0001); conversely Lower SpO_2_ at presentation increased risk of intubation. Female gender decreased risk of intubation (RR 0.34, 95% CI 0.13—0.92, *p* = 0.033); conversely male gender increased risk of intubation. Additionally, presence of delirium increased risk of intubation (RR 3.91, 95% CI 1.46—10.41, *p* = 0.006).

## Discussion

This study focused on a young and predominantly Hispanic COVID-19 patient population (92% of study population) in a predominantly Hispanic community (83%).^20^ Delirium tends to be more common in older patients, especially those with preexisting cognitive deficits.^7,8^ The population for this study was relatively small (total N of 133, N of 8 for patients with delirium), which limits the power and generalizability of the study. In this study population, 6% of patients were diagnosed with delirium, which was lower than expected. However, this may be explained by the convergence of several factors including the young subject population and established under-diagnosis of delirium by physicians and nurses. Lack of recognition of delirium is not unusual and happens for many reasons. One is the differing documentation used to describe it: altered mental status, confusion, encephalopathy, impaired consciousness, and acute brain failure among other terms.^21^ A study by Fogang et al. in Cameroon with a broader range of patient ages but a similar mean (54.6 years) reported a delirium rate of 5.6% and an impaired consciousness rate of 10.7%.^22^ While conducting this study we observed that many patients were described identically to the patients diagnosed with delirium (confused, agitated, inattentive, etc.), however the diagnosis of delirium was not documented. There was not a systematic delirium scale documented in the patients. There is a possibility that the sample of patients with delirium was larger than is reflected in this study which focused on patients with a delirium diagnosis in their medical record.

Previous studies have shown neurological manifestations such as delirium were found to be associated with more severe disease presentation and higher mortality rates.^23,24^ This suggests delirium is an indicator of a poorer prognosis in COVID-19 patients, which is consistent with previous studies though these studies tend to be oriented towards older patient populations.^3,16,17^ This study showed delirium was significantly associated with worse patient outcomes including longer hospital stay, longer ICU stay, higher rates of mechanical ventilation, more days on mechanical ventilation and higher mortality in this younger, predominantly Hispanic population. There was also a significant association between male gender, lower SpO_2_ and being on O_2_ therapy at presentation to ED with the diagnosis of delirium.

So, patients who initially presented worse at the ED in terms of O_2_ status were more likely to have a delirium diagnosis while in the hospital than those who presented with higher SpO_2_, and initially not on O_2_ therapy. Lower SpO_2_ at presentation was associated not only with delirium but also with 30-day mortality status and being placed on mechanical ventilation. This is in line with our finding of higher mortality rates and worse patient outcomes in those diagnosed with delirium, consistent with a study reported by Curyto et.al.^25^ Not only was a lower SpO_2_ at presentation associated with being placed on mechanical ventilation, but higher SpO_2_ at presentation had a significant relative risk of 0.96 (95% CI 0.95—0.97, *p* value of 0.022), which is equivalent to lower SpO_2_ having a higher relative risk of being placed on mechanical ventilation.

Worse initial presentation to the ED was significantly associated with worse patient outcomes aside from delirium: higher respiratory rate was associated with both 30-day mortality status and being placed on mechanical ventilation. Longer hospital stays, ICU stays, and more placement on and days with mechanical ventilation were associated with a higher 30-day mortality status as well as delirium. Our results are consistent with previous literature that indicates delirium is associated with worse health outcomes in COVID-19 patients.^16,22,24^

In terms of psychiatric history, patients diagnosed with delirium had pre-existing cognitive deficits and history of substance use. Cognitive deficits have been found in older patients to be associated with development of delirium, and we see here a confirmation of studies suggesting that association is also true in younger patients.^7,8^ Lower diastolic blood pressure was the only significant association found with presence of a psychiatric disorder. Presence of a psychiatric disorder was not associated with delirium diagnosis.

Other studies have noted an association between male gender and worse COVID-19 outcomes.^26,27^ Our data supports this conclusion as there was a significant association between male gender and the presence of delirium, higher mortality rates, and placement on mechanical ventilation. Similarly, increased age has traditionally been associated with higher COVID-19 mortality rates.^26^ Though this study focused on a younger demographic (≤65 years old), there was significant association between increased age and a 30-day mortality status of deceased.It is important to recognize the strengths and limitations of this study. This study is a retrospective chart review which introduces potential limitations and biases due to incomplete documentation, unrecorded information, variance of quality of information recorded, and issues of data abstraction. Additionally, there was no standardized method used to diagnose delirium in the clinical setting. This study is strong in its focus on young, Hispanic individuals which constitutes an under-studied community, and the rigor of the statistical analysis which revealed many significant associations and sources of increased risk.

## Conclusion

The mortality rate in COVID-19 patients with a delirium diagnosis was established in this cohort of adults aged 65 and younger, predominantly Hispanic patients to be higher than those without delirium. The presence of a pre-existing mental health diagnosis was not found to be a relevant co-morbidity in COVID-19 patients with a concurrent delirium diagnosis. We can suggest that clinical outcomes for those individuals with delirium are worse than their non-delirious counterparts. Individuals with delirium had longer, more involved hospital stays (ICU admission), were more often put on mechanical ventilation and had significantly higher mortality rates.

## Data Availability

All data produced in the present study are available upon reasonable request to the authors

## Notes

### Competing Interest Statement

The authors have declared no competing interest.

### Funding Statement

This study did not receive any funding

### Author Declarations

Institutional Review Board of TTUHSC El Paso waived approval for this work

